# Non-coding variants are a rare cause of recessive developmental disorders *in trans* with coding variants

**DOI:** 10.1101/2023.06.23.23291805

**Authors:** Jenny Lord, Carolina J Oquendo, Alexandra Martin-Geary, Alexander JM Blakes, Elena Arciero, Silvia Domcke, Anne-Marie Childs, Karen Low, Julia Rankin, Genomics England Research Consortium, Diana Baralle, Hilary C. Martin, Nicola Whiffin

## Abstract

**Purpose:** Identifying pathogenic non-coding variants in individuals with developmental disorders (DD) is challenging due to the large search space. It is common to find a single protein-altering variant in a recessive gene in DD patients, but the prevalence of pathogenic non-coding ;second hits; *in trans* with these is unknown.

**Methods:** In 4,073 genetically undiagnosed rare disease trio probands from the 100,000 Genomes project, we identified rare heterozygous loss-of-function (LoF) or ClinVar pathogenic variants in recessive DD-associated genes. Using stringent region-specific filtering, we identified rare non-coding variants on the other haplotype. Identified genes were clinically evaluated for phenotypic fit, and where possible, we performed functional testing using RNA-sequencing.

**Results:** We found 2,430 probands with one or more rare heterozygous pLoF or ClinVar pathogenic variants in recessive DD-associated genes, for a total of 3,761 proband-variant pairs. For 1,366 (36.3%) of these pairs, we identified at least one rare non-coding variant *in trans*. After stringent bioinformatic filtering and clinical review, five were determined to be a good clinical fit (in *ALMS1, NPHP3, LAMA2, IGHMBP2* and *GAA*).

**Conclusion:** We developed a pipeline to systematically identify and annotate compound heterozygous coding/non-coding genotypes. Using this approach we uncovered new diagnoses and conclude that this mechanism is a rare cause of DDs.

## Introduction

Large-scale exome or genome sequencing of individuals with developmental disorders (DDs) currently identifies a genetic diagnosis for ∼30-40% of individuals^1,2^. Analysis in the UK-based Deciphering Developmental Disorders (DDD) study has estimated that, of the ∼13,000 patients in that cohort, ∼41% are attributable to autosomal *de novo* coding mutations^3^, ∼7% to X-linked coding variants^4^ and ∼3% to autosomal recessive coding variants^5^. These calculations suggest that even after all DD-associated genes have been identified, a large fraction of individuals with DDs will not be attributable to a Mendelian-acting coding cause and hence remain genetically undiagnosed.

Variants in non-coding regions are increasingly being implicated in DDs. In DDD, it is estimated that ∼1% of DDs can be explained by *de novo* mutations in conserved regulatory elements^6^. In addition, variants in untranslated regions (UTRs)^7^ and deep intronic regions have been identified as causes of DD^8,9^. However, the overall contribution of Mendelian-acting non-coding variants to DDs has not been quantified. Sometimes, a single putatively deleterious coding variant is identified in a known recessive gene with a good match to an individual’s phenotype, without an obvious coding ‘second hit’ on the other haplotype. There are examples of non-coding second hits being identified in such patients, including deep intronic variants in individuals with respiratory disorders^10,11^. However, existing work has not systematically searched for this combined compound heterozygous coding/non-coding mechanism in large rare disease cohorts.

Here, we use genome sequence data from the Genomics England 100,000 Genomes project to investigate the contribution of inherited non-coding regulatory variants *in trans* with a deleterious coding variant to DDs. We identify individuals with a single loss-of-function or known pathogenic variant in a recessive DD gene, then systematically identify and annotate variants in nearby regulatory regions *in trans* that may constitute the ‘second hit’. We describe clinical follow-up on individuals whose phenotype was a potential fit to the identified gene, followed by transcriptomic investigation on one of them. Overall, we found that this combined compound heterozygous coding/non-coding mechanism explains a very small fraction of DDs but nonetheless accounts for clinically actionable diagnoses.

## Materials and methods

### Defining the candidate gene set

Genes within which variants are known to cause developmental disorders through a recessive mechanism were identified using the Developmental Disorders Gene to Phenotype (DDG2P) database (downloaded on 02/04/2019) as those with an ‘allelic requirement’ of ‘biallelic’ only (excluding those that also had other inheritance mechanisms), ‘mutation consequence’ including ‘loss of function’, and ‘DDD category’ of ‘confirmed’ or ‘probable’, resulting in a set of 793 candidate recessive genes (referred to henceforth as ‘DDG2P recessive genes’; Supplementary Table 1). We excluded the non-coding RNA gene *RMRP*.

### Identifying individuals with single coding variants

We used the Genomics England (GEL) 100,000 Genomes dataset (version 7). We only included probands recruited as full trios, comprising an affected proband and both unaffected parents, and that were aligned to GRCh38. We filtered out individuals with variants classified as either tier 1 or tier 2 in the GEL clinical filtering pipeline [https://re-docs.genomicsengland.co.uk/tiering/], which are most likely to have a monogenic diagnosis, plus any individuals with “solved” in their Exit Questionnaire, and probands with subsequently withdrawn consent up to v16 of GEL, leaving 4073 trios. In the remaining individuals, we searched for single heterozygous predicted loss-of-function (pLoF) variants in one of the 793 DDG2P recessive genes, defined based on annotations from Ensembl’s Variant Effect Predictor (VEP, v96)^12^ of “stop_gained”, “splice_acceptor”, “splice_donor”, and “frameshift”. pLoFs classified as low-confidence by LOFTEEv1.0 were excluded. Additionally, we identified single heterozygous variants in the DDG2P recessive genes that were annotated as pathogenic or likely pathogenic in ClinVar (CLNSIG of “Pathogenic”, “Likely_pathogenic”, or “Pathogenic/Likely_pathogenic”; downloaded on 21/09/21)^13^, with any predicted effect (i.e. not limiting to pLoF), and with a review status (CLNREVSTAT) of “criteria_provided,_multiple_submitters,_no_conflicts”, “reviewed_by_expert_panel”, or “practice_guideline”.

Variants were excluded for the following reasons: (1) variant not present in either parent (i.e. *de novo* variants); (2) allele frequency (AF) >0.5% across the 4073 included trios; (3) popmax AF >0.5% gnomAD v2.1.1^14^; (4) <25% or >75% of sequencing reads at that position in the proband contain the variant; (5) VCF “Filter” not “PASS”; and (6) sequencing depth at variant position <6.

## Defining non-coding regulatory regions

For each of the 793 DDG2P recessive genes, we defined the coordinates of all intronic regions, the 5’UTR and 3’UTRs, and a candidate upstream promoter region comprising the first 5,000 bps directly upstream of the transcription start site (TSS). Regions were identified for all MANE v1.0 transcripts (MANE Select and Plus Clinical)^15^. Upstream promoter regions were sub-divided into a ‘core promoter’, comprising the first 200 bps upstream, and an ‘extended promoter’ as the remaining region.

Additionally, since most DD patients have an abnormality of the nervous system, we used regions of DNA accessible in fetal brain that were identified using sci-ATAC-seq^16^. We filtered identified peaks to only those identified in ≥5% of cells from fetal cerebrum, or that were in the top 5% of cell-type specificity scores. Peaks were further filtered to only those overlapping the defined upstream promoter region of a DDG2P recessive gene, or that are identified as co-accessible with a region fitting this promoter overlap criterion^16^.

Genomic coordinates of all candidate regions are in Supplementary Table 2.

## Identifying candidate non-coding ‘second hit’ variants

For each proband-variant pair (i.e. combination of proband and single identified pLoF or ClinVar variant), candidate second hit variants were identified across the non-coding regions defined for the gene containing the coding variant. Only “PASS” variants in the VCF with <25% or >75% of sequencing reads at that position containing the variant were considered. Only heterozygous variants transmitted by the alternative parent to the coding variant (i.e. following expected recessive inheritance), with gnomAD v3.0 filtering allele frequency (FAF)^14^ ≤ 0.5% across all major continental populations, and internal allele frequency in GEL ≤ 0.5% (calculated from the aggregated multi-sample VCF) were retained. Variants were removed if they overlapped the coding sequence of any MANE transcript (n=12) or had a ClinVar annotation of ‘Benign’, ‘Likely Benign’ or ‘Benign/Likely Benign’ (n=13). This gave 1,366 proband-variant pairs with both a single coding variant plus a non-coding variant *in trans*.

The non-coding variants were prioritised if they matched any of the following region-specific annotations, and the prioritised proband-variant pairs were subsequently subjected to manual clinical review:

- Intronic variants with SpliceAI ≥0.1 (including UTR introns)
- Promoter variants in the ‘core’ region (the first 200bps directly upstream of the TSS), or that overlap either a sci-ATAC region or a transcription factor binding site annotation from GreenDB^17^ and have CADD ≥15
- 5’UTR variants with SpliceAI ≥0.1, overlapping a transcription factor binding site annotation from GreenDB^17^, with an annotation from UTRannotator^18^, or within an internal ribosome entry site (IRES) from IRESbase^19^
- 3’UTR variants with SpliceAI ≥0.1, overlapping an experimental miRNA binding site collated from four studies^20–23^, or within a polyadenylation signal sequence (defined using Gencode PolyA feature annotation or within a canonical AATAAA motif).
- A variant in any region with PhyloP ≥5, and/or CADD ≥20

## Clinical filtering

Individuals with candidate non-coding second hit variants were manually reviewed to assess whether the gene was a good clinical match for their phenotype. The individual’s phenotype terms and the disease class under which they were recruited were reviewed by a consultant clinical geneticist [D.B.], against the expected presentation of biallelic pathogenic variants in the relevant genes. Using information on disease presentation from OMIM^24^, DECIPHER^25^, and expert knowledge, each proband-gene pair was classified as “probable”, “possible” or “unlikely”.

Where there was a “probable” match between the proband;s recorded phenotype and that associated with the gene, clinical contact forms were submitted to GEL. Clinicians who responded were asked to review the gene as a potential diagnostic candidate for their patient. Where a plausible phenotypic match was confirmed, the clinician was invited to offer the patient RNA profiling. In one instance (*NPHP3*), the variants had already been recorded as a “partial diagnosis” by the proband’s recruiting centre, so contact was not initiated.

Genes were annotated with whether or not they were classified as ‘Green’ in the GEL PanelApp resource^26^ in a gene panel that was applied to the participant, to flag genes that had been considered *a priori* a possible cause of the participant’s phenotype.

## RNA-sequencing

The participant gave informed consent to undergo RNAseq under the University of Southampton’s Splicing and Disease Study, with ethical approval from the Health Research Authority (IRAS ID 49685, REC 11/SC/0269) and the University of Southampton (ERGO ID 23056). Whole blood samples were collected in PAXgene Blood RNA tubes and RNA was extracted using the PAXgene Blood RNA kit (PreAnalytiX, Switzerland). Random hexamer primers were used to generate complementary DNA (cDNA) via reverse transcription.

RNA libraries were prepared by Novogene (Hong Kong) with rRNA and globin depletion (NEBNext kits), using the NEBNext Ultra Directional RNA Library Prep Kit for Illumina (New England Biolabs, MA). Sequencing (also by Novogene) was conducted to generate at least 70 million 150bp paired-end reads on the HiSeq 2000 (Illumina, CA). Raw read quality filtering and adapter trimming were performed by Novogene.

Reads were aligned to the reference genome (GRCh38) using STAR (v2.6.1c) and sequencing reads and sashimi plots in the vicinity of the variants were manually assessed using the Integrative Genomics Viewer (IGV, Broad Institute, MA). rMATS-turbo v4.1.2^27^ was used to detect aberrant splicing, with results filtered to remove events that were outliers in multiple individuals, and OUTRIDER^28^ to test for expression outliers. Twenty-nine additional patients with diverse phenotypes recruited to the Splicing and Disease study and sequenced in the same batch were used as controls. ggsashimi^29^ was used to generate sashimi plots to visualise splicing events. Intron coverage was calculated for all samples in the sequencing batch using HTSeq^30^, normalised by total gene read count (from STAR) and visualised using ggplot2^31^ in R version 3.5.1^32^.

## Results

### Identifying and filtering candidate non-coding second hits in genetically undiagnosed rare disease probands

We identified 4,073 rare disease trio probands in GEL without an existing genetic diagnosis. These were recruited for a wide range of primary phenotypes, of which the most common was Neurology and Neurodevelopmental Disorders (1,711 probands; Supplementary Figure 1). In 2,430 of the 4,073 probands (59.7%), we found a single heterozygous pLoF or ClinVar (likely) pathogenic variant in one of 793 DDG2P recessive genes. 940 probands had multiple such variants in different genes (Figure 1a), giving a total of 3,761 proband-variant pairs, including 2,574 pLoFs and 1,187 additional ClinVar variants.

**Figure 1:**
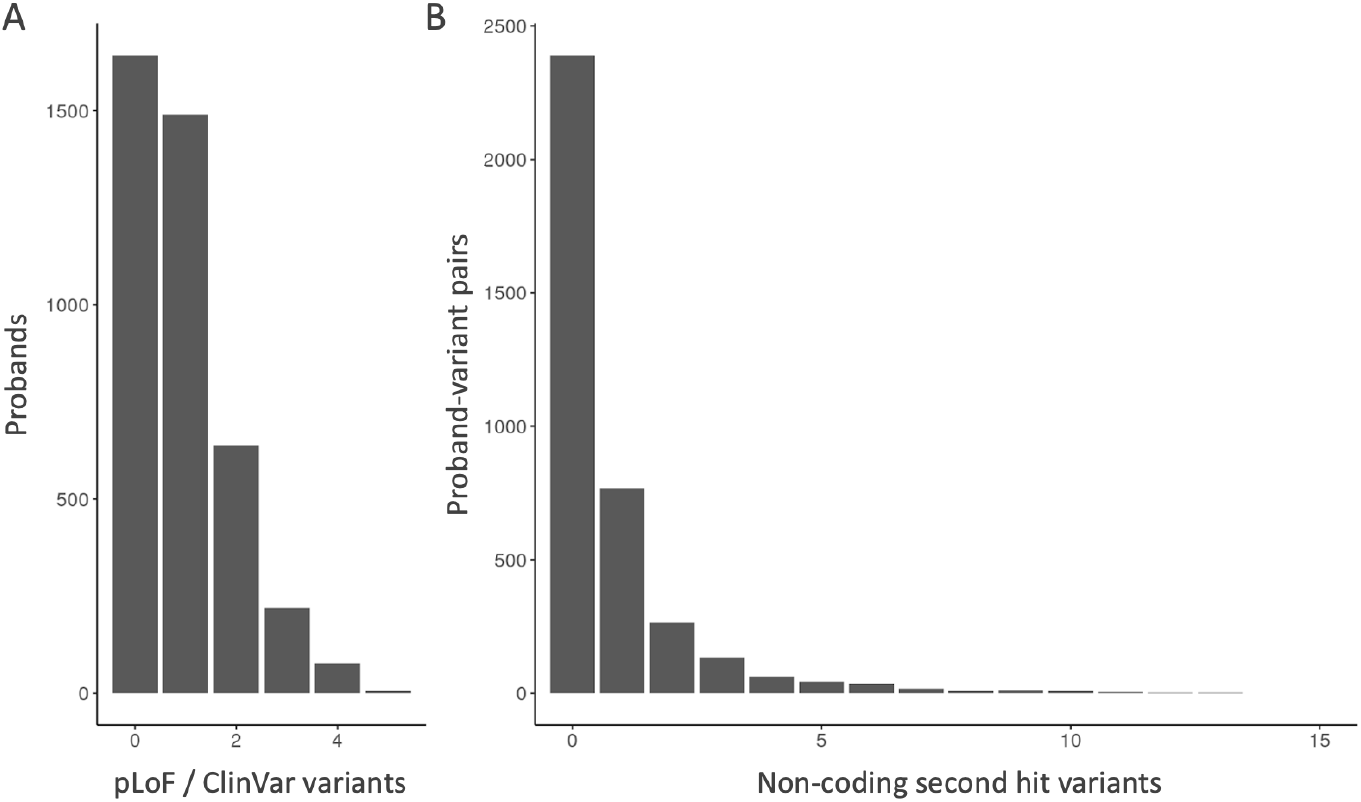
The number of candidate variants identified per proband. (a) Bar plot of the number of single pLoF and/or ClinVar (likely) pathogenic variants per individual; (b) Bar plot of the number of candidate non-coding second hit variants per proband-variant pair. The x-axis is truncated at 15. Four proband-variant pairs had >15 non-coding second hit variants with counts of 16, 20, 25 and 38.

We defined 16,847 distinct non-coding regions associated with our 793 DDG2P recessive genes (Supplementary Table 2), spanning on average 85,937 bps for each gene (range: 5,624 to 2,305,299 bps; Supplementary Table 3). For 1,366 (36.3%) of our proband-variant pairs, we identified at least one rare non-coding variant *in trans* (i.e. inherited from the alternate parent to the coding variant) that passed our quality filters. 597 proband-variant pairs had more than one candidate second hit variant (Figure 1b), giving a total of 2,973 proband-variant-second hit combinations. The vast majority of our candidate second hit variants were intronic (2,744; 92.3%), reflecting the composition of our non-coding search space.

Given our expectation that most non-coding region variants have a very small, if any, regulatory impact, we created a stringent set of filters to narrow down our list of candidate second hit variants to those that seemed most likely to have an effect (see Methods). After filtering, we retained 52 candidate second hit variants in 52 probands: thirty-five intronic, eight in the promoter region, six in the 5’UTR, and three in the 3’UTR (Figure 2).

**Figure 2:**
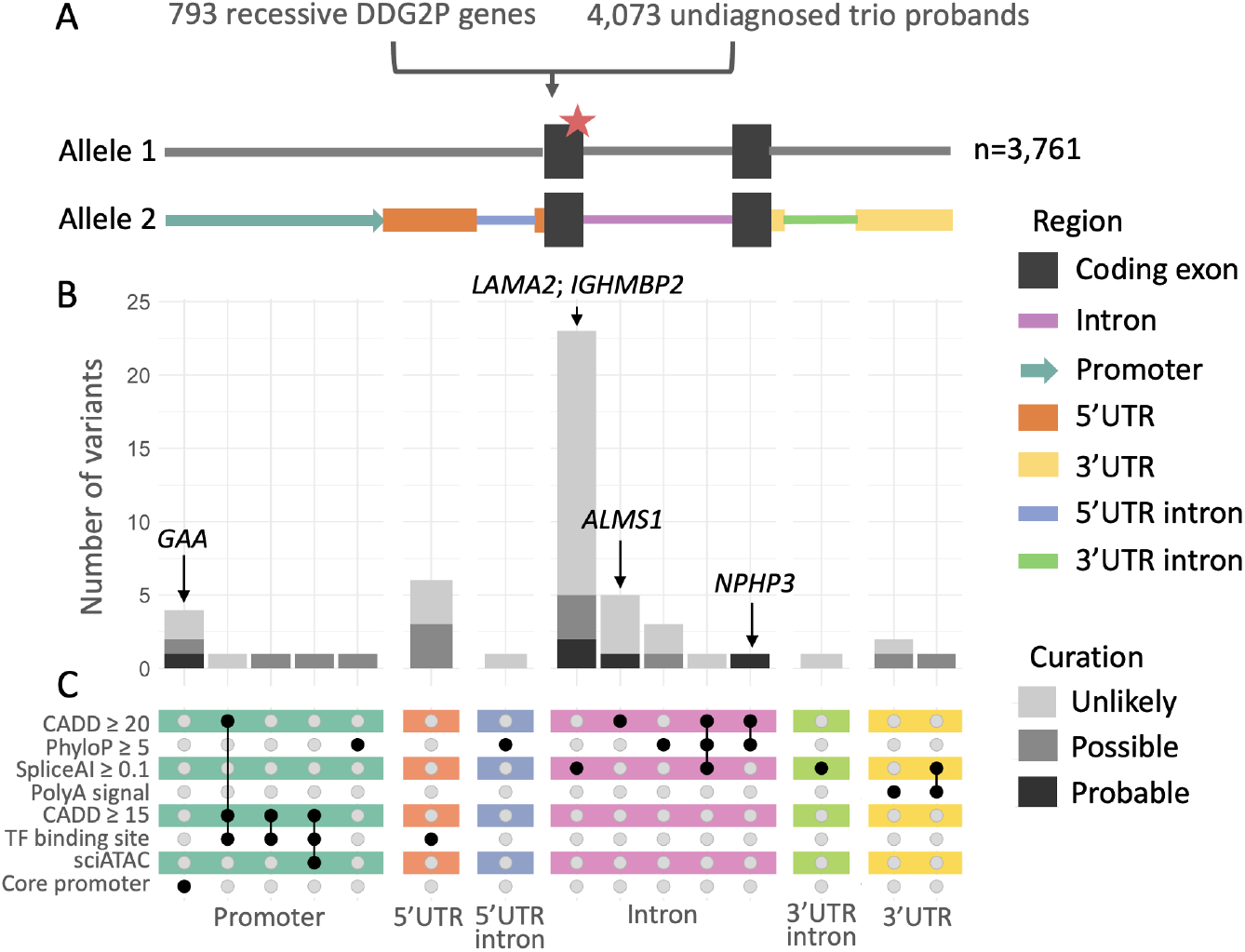
An overview of our approach and the distribution of 52 candidate non-coding second hits across different regions. (A) Details of our search focusing on 793 genes with a single pLoF or ClinVar (Likely) Pathogenic variant in 4,073 undiagnosed trio probands. The different candidate non-coding regions analysed for a second hit are shown in colour; (B) Count of candidate variants clinically curated as ‘probable’, ‘possible’ or ‘unlikely;, split by region and annotations. The indicated genes are those that were assessed to be a ‘probable’ fit. (C) Upset plot of region-specific annotations used to prioritise candidate variants.

### Assessing candidate variant match to clinical phenotype

Our 52 candidate coding/non-coding variant pairs were manually reviewed to assess whether they represented a credible explanation for each proband’s phenotype. Five (9.6%) of the variants were classified as a “probable” match (Table 1), thirteen (25.0%) as “possible”, and the remainder as “unlikely”. Of the five that were classified as “probable”, all except *NPHP3* were “green genes” on relevant panels that were applied to the proband by GEL^33^ (i.e. had been considered a plausible cause *a priori*), compared to only twelve of the remaining forty-seven (25.6%; odds ratio = 11; Fisher’s *P*=0.02).

**Table 1:** Candidate coding/non-coding variant pairs. Shown are variant details, selected annotations, and phenotypic data relating to the proband. HPO: Human Phenotype Ontology; AF: Allele Frequency; FAF: gnomAD filtering AF.

| Gene; Transcript | Phenotype |  | Coding variant | Non-coding region variant |  |  |  |  |  |
| --- | --- | --- | --- | --- | --- | --- | --- | --- | --- |
|  | Normalised specific disease | Abstracted selected HPO terms |  | Details | gnomAD FAF | GEL AF | SpliceAI | PhyloP | CADD |
| GAA;<br>ENST00000302262 | Limb girdle muscular dystrophy | Abnormality of the calf musculature; muscular dystrophy; respiratory insufficiency; Abnormality of the eye; progressive muscle weakness | chr17:80118288:G:A Nonsense | chr17:80101399:C:G Core Promoter | 2.75x10<sup>-3</sup> | 3.17x10<sup>-3</sup> | NA | -0.19 | 5.88 |
| NPHP3;<br>ENST00000337331 | Proteinuric renal disease | Abnormal renal corpuscle morphology; abnormal liver morphology; abnormal urine metabolite level | chr3:132691199:G:A Nonsense | chr3:132684549:C:T Intronic | 5.14x10<sup>-6</sup> | 8.95x10<sup>-5</sup> | 0.04 | 6.15 | 21 |
| ALMS1;<br>ENST00000613296 | Cone dysfunction syndrome | Abnormal visual electrophysiology; Abnormal eye physiology; Abnormal retinal morphology; Abnormality of vision | chr2:73572648:AC:A Frameshift | chr2:73573562:G:A Intronic | 1.72x10<sup>-3</sup> | 1.73x10<sup>-3</sup> | 0.01 | 3.84 | 20.2 |
| LAMA2;<br>ENST00000421865 | Congenital myopathy | Abnormal skeletal muscle morphology; muscle weakness; abnormal muscle physiology; abnormal joint physiology | chr6:129316089:C:T Nonsense | chr6:129475360:G:GT Intronic | 4.71x10<sup>-4</sup> | 5.88x10<sup>-4</sup> | 0.1 | NA | 8.48 |
| IGHMBP2;<br>ENST00000255078 | Charcot-Marie-Tooth disease | Peripheral axonal degeneration | chr11:68936904:GC:G Frameshift | chr11:68929807:G:A Intronic | 9.51x10<sup>-5</sup> | 1.66x10<sup>-4</sup> | 0.12 | -1.91 | 0.21 |

One of the “probable” cases, with a frameshift variant and an intronic variant in *ALMS1* 23bp from a splice donor site, also had a two-exon duplication *in cis* with the intronic variant. This had already been considered as a potential diagnosis by the recruiting site, with the pLoF variant classified as pathogenic and the duplication as a VUS. Additionally, the intronic variant we detected was present in a homozygous state in three members of the UK BioBank (https://decaf.decode.com/region/chr2:73573557-73573567). Taken together, we felt this meant the intronic variant was unlikely to be pathogenic.

For the remaining four probable diagnoses, we attempted to contact the proband’s recruiting clinician through the GEL portal. We discuss each of these cases below.

### GAA

We identified a variant in the promoter of *GAA*, 182 bps upstream of the transcription start site (TSS) (GRCh38; chr17:80101399:C:G) *in trans* with a nonsense variant (GRCh38; chr17:80118288:G:A) in a proband with a phenotype most similar to limb girdle muscular dystrophy (LGMD). Mutations in *GAA* cause a glycogen storage disorder called Pompe disease^34,35^. *In silico* scores suggest this variant is unlikely to be deleterious (PhyloP <0; CADD=5.9), but following contact with the clinical team, biochemical assays confirmed marked deficiency of GAA enzymatic activity in the patient and hence a diagnosis of Pompe disease. Of note, this patient had had GAA enzymatic testing undertaken several years previously and, at that stage, was within normal limits. Across the full GEL cohort (i.e. not limited to trios), we observed this variant in a total of six probands who also carried a rare missense or pLoF variant (phase unchecked). Three of these probands, including our initial case, were reported to have LGMD.

Subsequently, we found that all three GEL LGMD probands with the promoter variant also carried a 5’UTR intronic variant, -32-13T>G (GRCh38; chr17:80104542:T:G). This variant is reported as pathogenic in 45 submissions to ClinVar (variation ID:4027), is observed in 36-90% of Pompe disease cases (tending to cause later-onset disease), and has been demonstrated to impact splicing, albeit with an incomplete/leaky effect^36,37^. The promoter and 5’UTR intronic variants were confirmed to be *in cis* in our index proband. The 5’UTR intronic variant was missed in our initial search for second hit variants due to its high frequency in gnomAD (maximum AF 0.0073 in Latinos/Admixed Americans).

Whole blood based RNAseq was conducted on the index proband to assess the impact of the variants on gene expression and splicing. The nonsense variant would be expected to result in nonsense-mediated decay (NMD) of transcripts from that allele, and whilst OUTRIDER^28^ did not detect lower expression of the gene relative to other patients in the same sequencing batch, manual inspection of reads in IGV showed a lower proportion of reads carrying the alternate allele (19 vs 46, 29%, Figure 3a). The normal expression level of the gene suggests that the promoter variant does not impact gene expression.

**Figure 3:**
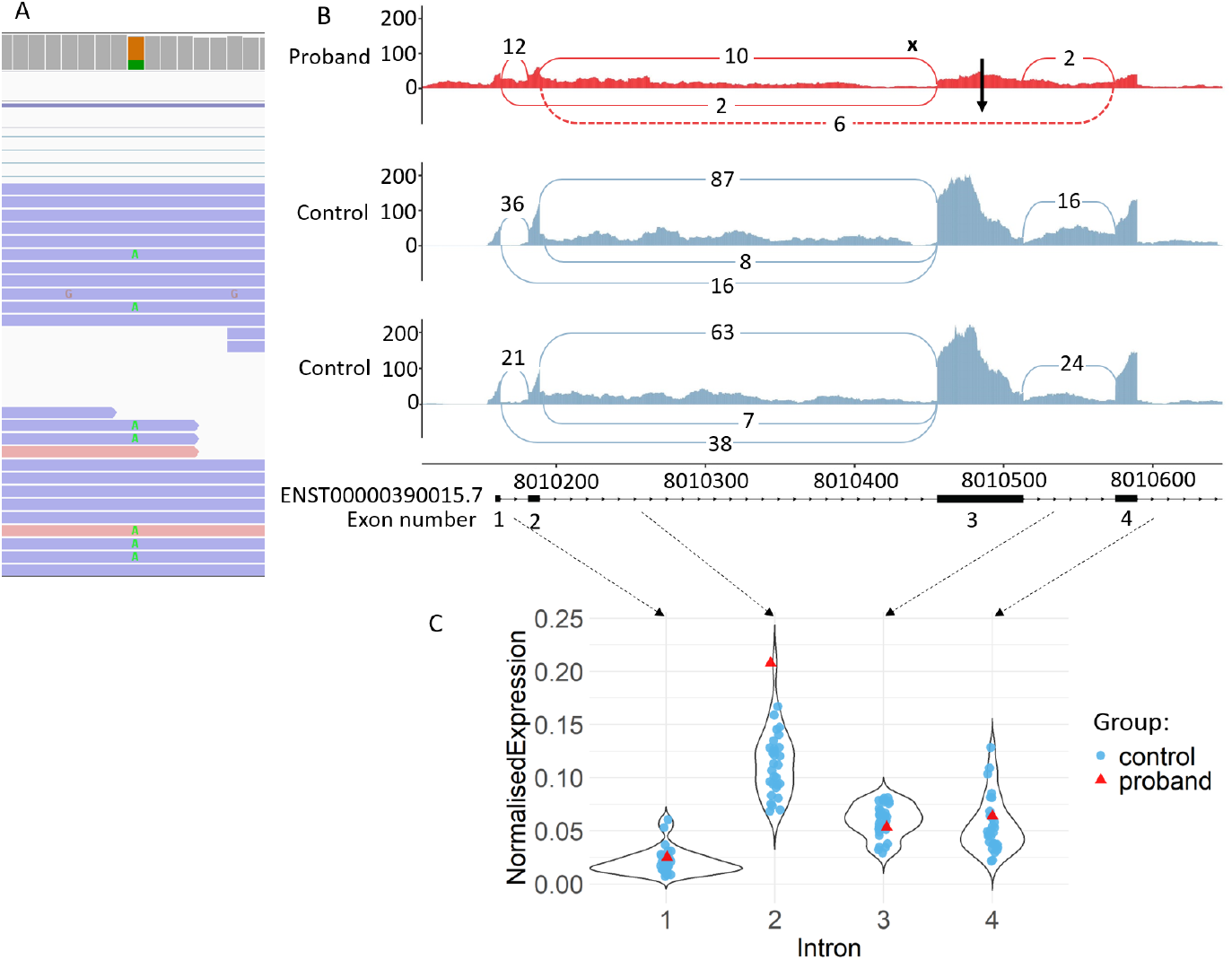
RNAseq from the proband with GAA variants. A) RNAseq reads covering nonsense variant chr17:80118288:G:A. 46 reads carry the reference allele while 19 carry the alternative allele. B) Sashimi plot showing splicing in the proband plus two controls from the same sequencing batch. Skipping of exon 3 (ENST0000390015.7) is observed in the proband but not in controls (dashed line, black arrow) due to the variant 13bp upstream of the exon 3 splice acceptor site (black “x”). C) Normalised expression level per intron for the proband (red) plus 29 controls (grey) sequenced in the same batch. Proband has higher intronic coverage than all controls for intron 2, suggesting intron retention may be occurring.

However, we detected altered splicing patterns at the 5’ end of *GAA* near the 5’UTR intronic variant using rMATS (Supplementary Table 4), showing skipping of exon 3 (ENST0000390015.7, Figure 3b), which was not observed in controls, and has previously been reported as an outcome of the -32-13T>G variant^38^. This exon contains the start of the *GAA* protein coding sequence, and skipping of this exon would remove the first 19% of the protein (182 of 952 amino acids). Additionally, we observed a large number of reads mapping to the intronic regions in the area around the 5’UTR variant, suggesting there may be full retention of introns, particularly intron 2 (Figure 3c).

Together, these investigations strongly imply that the -32-13T>G splice-altering variant is more likely to be the functional non-coding second hit in *GAA* than the promoter variant we initially identified.

### NPHP3

In a proband with proteinuric renal disease, we found two variants in *NPHP3*, which is known to cause nephronophthisis^39^. These were a nonsense variant in exon 18 (GRCh38; chr3:132691199:G:A), reported to be pathogenic in ClinVar with multiple submitters (variation ID:571559), and an intronic variant (GRCh38; chr3:132684549:C:T) 5bp from the splice donor site of exon 24. Although the SpliceAI score for this variant is below our threshold (0.04), donor +5 variants are known to be under strong selective constraint and often impact splicing^40,41^. The high CADD (21) and PhloP (6.5) scores for this variant are supportive of a deleterious impact. Abolition of this splice donor site would lead to the addition of four amino acids before an inframe stop codon, which would be predicted to lead to NMD.

This pair of variants had been triaged by GEL’s tiering pipeline and classified as “Tier 3” variants, and had already been assessed as a potential diagnosis by the proband’s recruiting centre. The stop gained variant had been classified as “pathogenic”, and the near-splice variant as “likely pathogenic”, with the overall assessment that the variant pair was partially responsible for the patient’s phenotype.

### LAMA2

In a proband reported to have congenital myopathy, we found a nonsense variant (GRCh38; chr6:129316089:C:T) previously reported to be pathogenic in eight ClinVar submissions (variation ID:92956), plus an intronic 1bp insertion (GRCh38; chr6:129475360:G:GT) 30bp from the splice acceptor site of exon 53, (ENST00000421865.3). SpliceAI predicts loss of the nearby acceptor (0.05) and of the donor of the same exon 41bp away (0.1), predicting an exon skipping impact. Variants that disrupt the canonical acceptor site of exon 53 have previously been reported in patients with muscular dystrophy (ClinVar variation IDs 1068380 and 954079)^42^.

Biallelic variants in *LAMA2* cause muscular dystrophies, with a correlation between phenotype and genotype reported^43^. Biallelic truncating variants in *LAMA2* are associated with a severe, early onset congenital muscular dystrophy type 1A (MDC1A), while missense and some splicing variants lead to a less severe and often later onset LGMD. The proband has a relatively static phenotype with reasonably well preserved muscle function, which would be consistent with the potentially leaky skipping of the small (12bp), in-frame exon. The patient’s recruiting clinician confirmed the variant pair as a plausible diagnosis. Demonstration that the intronic variant does impact splicing via RNA investigations are needed to confirm this, but we have been unable to obtain RNA.

### IGHMBP2

A pair of variants in *IGHMBP2*, a gene known to cause Charcot-Marie-Tooth disease (CMT), were found in a proband reported to have CMT disease. We identified a 1bp insertion in exon 13 (GRCh38; chr11:68936904:GC:G. ENST00000255078.8), expected to lead to a frameshift *in trans* with an intronic SNV (GRCh38; chr11:68929807:G:A), 450bp from the splice donor site of exon 8. This variant has previously been reported in ClinVar as a VUS (variation ID:2430634), with an accompanying comment asserting functional testing of the variant revealed leaky abnormal splicing. The variant introduces a new “AG” motif, with the potential to act as a splice acceptor site (SpliceAI acceptor gain 0.12), which would likely result in the generation of a cryptic exon. RNA studies are needed to confirm this impact, but we have been unable to obtain RNA.

Again, there is reported genotype-phenotype correlation for this gene, with the severity of the disorder linked to the nature of the variants. Loss-of-function variants and missense variants in conserved residues in or near the DNA helicase domain are associated with severe, distal hereditary motor neuronopathy (type VI), while axonal CMT (type 2S) is associated with less disruptive variants which allow some residual protein function^44^. This milder CMT phenotype would be expected if the intronic variant;s impact on splicing is incomplete.

## Discussion

Here, we systematically identified, annotated, and prioritised non-coding variants *in trans* with high-impact or known pathogenic coding variants, across a large cohort of undiagnosed individuals with rare disease. We successfully identified likely diagnoses for six probands, three with the same non-coding second hit in *GAA*, and three further probands each with unique variant combinations in distinct genes.

A key conclusion of this work is that the proposed mechanism of a non-coding second hit in combination with a single heterozygous coding variant is unlikely to explain a large fraction of undiagnosed DD cases. This is despite a large fraction (∼60%) having a single pLoF in a known recessive DD gene. Nevertheless, there are some key reasons why our reported diagnostic rate is likely an underestimate. Firstly, we did not perform an upstream clinical review due to the broad spectrum of DDs and incompleteness of phenotypic data in GEL, but rather included all rare disease probands without a complete existing diagnosis. Hence, not all of the 4073 tested probands have a phenotype compatible with our DD gene list. Secondly, we focussed our search for non-coding variants in regions within (introns and UTRs) or with clear links to genes (directly upstream), defined using a single representative transcript per gene. This approach will exclude many regulatory elements, including more distal enhancer regions, although arguably captures the regions most likely to contain variants of large effect. Thirdly, we focused on SNVs and small indels, so will have missed non-coding structural variants (such as the most likely second hit in *ALMS1* which was an exonic deletion). Fourthly, it was necessary to perform very strict filtering of candidate non-coding variants to reduce the number for clinical review, as a substantial proportion of our proband-variant pairs (∼36%) had at least one non-coding variant *in trans*. However, we are still lacking knowledge of the ‘regulatory code’ and tools to effectively filter non-coding region variants. Our stringent region-specific filtering approach could likely be improved as our knowledge of non-coding region variants and their impacts in disease continues to evolve. Fiftly, it is likely that some individuals with non-coding second hits will have already been diagnosed by GEL and hence not have been included in our initial cohort. These would include probands with splice region variants that were flagged as tier 1 or 2, or individuals for which the recruiting centre looked at tier 3 variants (potentially because local testing flagged a single hit in a candidate recessive gene). Finally, due to the difficulty of re-contacting recruiting clinicians through the GEL framework, we limited our follow-up efforts to only individuals where our initial clinical review suggested a variant pair as the ‘probable’ cause of the reported phenotype. It is likely that some of the variants/genes classified as ‘possible’ are also *bona fide* diagnoses, especially given the phenotypic information within GEL is often incomplete.

A large proportion of our identified non-coding second hits are intronic. This is expected, given that the vast majority of the search-space per gene (∼90%) is intronic. It is also relevant to note that DD recessive genes on average have much shorter 5’UTRs than the average across all genes (and indeed their dominant counterparts), reflecting the lower importance of translational regulation in these genes^45^. We would therefore not expect to find many high-impact 5’UTR variants across our gene set.

Our analyses, along with prior reports, suggest that the *GAA* 5’UTR -32-13T>G splice-altering variant is a more likely damaging second hit than the initially identified promoter variant *in cis*. This 5’UTR variant was not picked up in our filtering pipeline as it was over our allele frequency threshold. This underscores the potential for hypomorphic variants, i.e. those that are not complete loss-of-function, to be found at higher frequencies in the population. To our knowledge, this is the first time RNAseq has been conducted on a patient with the common 5’UTR -32-13T>G *GAA* variant. We confirmed the previously reported skipping of exon 3, which contains the start codon, and found evidence of full intron retention that has not previously been reported, likely due to differences in methodology. Prior validations had utilised RT-PCR or mini-gene assays, neither of which would be expected to detect this intron retention event, due to the much larger size of the amplicon, or the lack of native context used. RNAseq of other probands with the variant, particularly long read sequencing, would help clarify the exact nature of the aberrant transcripts generated. Larger-scale missplicing events, such as full intron retention and multi-exon skipping have been reported to be less frequent than single exon skipping or cryptic splice site usage^46^. With increasing use of RNAseq, more of these larger scale events may be detected, potentially revealing them to be more common than previously appreciated. This may necessitate the reclassification of variants previously believed not to affect splicing, but whose effects were not adequately captured by previous methodologies.

Our results have important implications for genetic testing guidelines and consideration of the strength of evidence assigned to variants found *in trans* with pathogenic coding variants (i.e. activation of PM3 in the ACMG/AMP framework^47^). Despite the increase in search-space when including non-coding regulatory region variants, with a careful filtering approach the number of candidate non-coding second hits can be kept to a manageable level, and hence PM3 can still be applied at a moderate strength^48^.

In summary, we have developed a systematic approach to identify non-coding second hits in recessive genes, highlighting how damaging non-coding variants can be annotated to find new diagnoses for undiagnosed DDs. Through this, we conclude that this mechanism is unlikely to account for a large proportion of missing DD diagnoses. Future work should couple RNAseq with genome sequencing to attempt to identify additional pathogenic non-coding variants, but should also consider other potential explanations for undiagnosed patients, such as coding variants in novel genes^3^ as well as oligogenic and polygenic contributions^49^.

## Data Availability

Data were analysed inside the secure Genomics England research environment. All sharable data were exported with approval and are included in the manuscript or as supplementary tables. Supplementary Table 2 is hosted on GitHub due to its size: https://github.com/Computational-Rare-Disease-Genomics-WHG/non-coding_second_hits

## Acknowledgements

JL is supported by a University of Southampton Anniversary Fellowship. CJO, JL and DB are supported by NIHR Research Professorship to DB (RP-2016-07-011). KL is supported by a NIHR Doctoral Research Fellowship (NIHR302303). HCM is supported by Wellcome grant 220540/Z/20/A to the Sanger Institute. NW and AM-G are supported by a Sir Henry Dale Fellowship to NW, jointly funded by the Wellcome Trust and the Royal Society (220134/Z/20/Z) and research grant funding from the Rosetrees Trust (PGL19-2/10025). AB is supported by a Wellcome PhD Training Fellowship for Clinicians.

This research was made possible through access to the data and findings generated by the 100,000 Genomes Project. The 100,000 Genomes Project is managed by Genomics England Limited (a wholly owned company of the Department of Health and Social Care). The 100,000 Genomes Project is funded by the National Institute for Health Research and NHS England. The Wellcome Trust, Cancer Research UK and the Medical Research Council have also funded research infrastructure. The 100,000 Genomes Project uses data provided by patients and collected by the National Health Service as part of their care and support.

EA is a current employee of Flagship Labs 86, Flagship Pioneering. All other authors declare no conflicts of interest.

This research was funded in whole, or in part, by the Wellcome Trust. For the purpose of open access, the author has applied a CC BY public copyright licence to any Author Accepted Manuscript version arising from this submission.

## Supplementary Data

### Supplementary Tables

Supplementary Table 1: List of DDG2P recessive genes and MANE Select v1.0 transcripts Supplementary Table 2: List of annotated non-coding regions for each DDG2P recessive gene.

Supplementary Table 3: Overview of the non-coding search space. Shown is the total search-space across all transcripts corresponding to each region type. Note, a base could be counted more than once if it has different annotations for different genes. The mean, min and max are calculated for the number of genes with at least a single base assigned to that region.

Supplementary Table 4: rMATS differential splicing results for proband with GAA variants. Information on the differential splicing event associated with the GAA UTR variant, including coordinates of exons, counts of reads supporting the skipping and inclusion events, FDR corrected p-value and inclusion level difference between the proband and the controls.

## Supplementary Figures

**Supplementary Figure 1:**
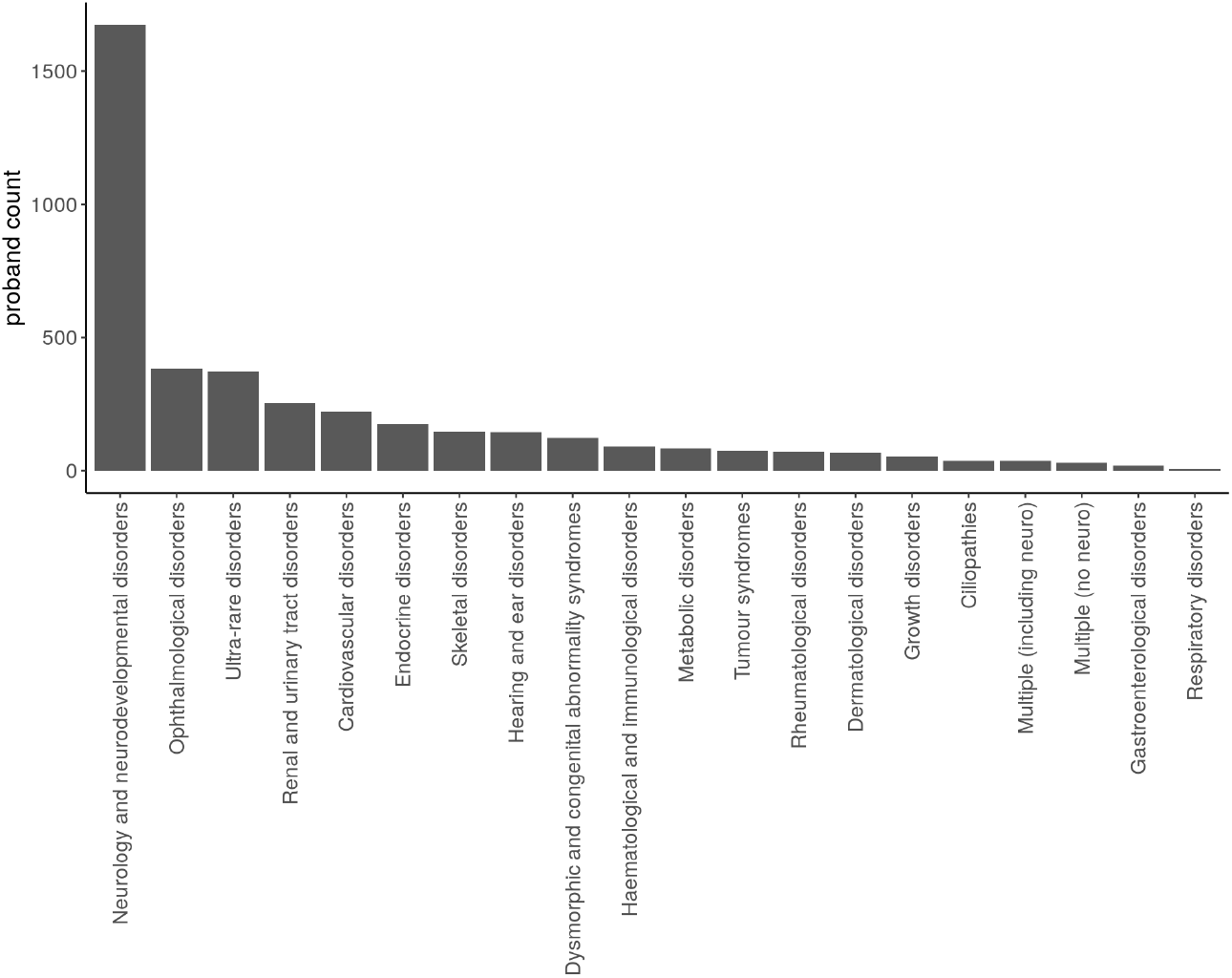
Listed ‘normalised disease group’ for the initial set of 4,073 undiagnosed probands. 1,711 (42%) have “Neurology and neurodevelopmental disorders” as a listed normalised disease group. Due to Genomics England rules, disease groups containing fewer than five probands have been removed.

## Notes

### Author Declarations

The 100,000 Genomes Project Protocol has ethical approval from the HRA Committee East of England Cambridge South (REC Ref 14/EE/1112). This study was registered with Genomics England under Research Registry Projects 155. The University of Southampton Splicing and Disease Study has ethical approval from the Health Research Authority (IRAS ID 49685, REC 11/SC/0269) and the University of Southampton (ERGO ID 23056).

